# Optimisation of a cervical cancer screening model based on self-sampling for human papillomavirus testing

**DOI:** 10.1101/2024.10.16.24315599

**Authors:** María Besó Delgado, Josefa Ibáñez Cabanell, Susana Castán Cameo, José Joaquín Mira Solves, Mercedes Guilabert Mora, Mercedes Vanaclocha Espí, Marina Pinto Carbó, Dolores Salas Trejo, Oscar Zurriaga Llorens, Ana Molina-Barceló

## Abstract

The use of self-sampling devices in population-based cervical cancer screening programmes (CCSP) is both an opportunity and a challenge in terms of implementation.

**Objective:** To understand the expectations, preferences, and difficulties perceived by women as regards different screening models and self-sampling devices, with the aim of identifying strategies to achieve high CCSP participation rates.

**Methods:** This study is based on qualitative research techniques, consisting of an individual interview using a questionnaire with semi-structured questions, followed by a group interview. Sessions were held simultaneously with 4 groups (7-8 women aged between 35 and 65). Women assessed various aspects of the programme (information dissemination, invitation, receipt of results, etc.) and five self-sampling devices.

**Results:** If screening were carried out via self-sampling, 96.4% of women said they would take the test. Most women preferred to receive information on the CCSP or return their samples at their health centres (86.2% and 86.2%), and the most popular method for receiving both the programme invitation and results is by SMS (58.6%, 65.5%).

Simplicity and ease of use are the key features of the device accepted by the largest number of women, the FLOQSwab. Another highly rated feature is the attractive design of the Evalyn Brush, as this was the preferred device of the largest number of women.

The existence of other screening programmes that use self-sampling devices (the colorectal cancer prevention programme) is an opportunity as regards acceptance of this new programme. Some women are unsure of how to use the devices correctly.

A large number of women accept self-sampling and reveals significant differences in the degree of acceptance of different self-sampling devices. Selecting the most accepted device is key to achieving high CCSP participation rates, and these programmes should be accompanied by adapted information campaigns to reach the most vulnerable groups and ensure equity.

## Introduction

Cervical cancer is the fourth most common cancer affecting women in terms of both incidence and mortality, with an estimated global mortality rate of 7.3 per 100,000 per year in 2020, according to the International Agency for Research on Cancer (IARC)[1]. In most cases, these cancers are secondary to persistent high-risk human papillomavirus (HPV) infection.

Carcinogenesis is a multi-step, long-term process in which both genetic and morphological changes occur in the cells of the cervix. As population-based cervical cancer screening programmes are widely proven to be costeffective and make it possible to detect and treat lesions at an early stage, their implementation is included in European Commission recommendations [2-3] and they are a fundamental tool for preventing cervical cancer. However, many European countries, such as Spain, have opportunistic cervical cancer screening programmes [4-5] that reach just a small section of the target population and achieve limited benefits, while also generating inequalities. Countries face the challenge of converting these opportunistic programmes into population-based programmes.

Moreover, in recent years the recommendations of this screening programme have been modified. Firstly, HPV detection has been incorporated as a primary screening test to replace conventional cervical cytology testing, as recommended by numerous organisations and agencies [2, 6, 7, 8]. With the acceptance and approval of this new primary test, the use of self-sampling devices, which allow women to collect an HPV testing sample at home, has been researched and approved [9]. The effectiveness of these tests has been widely demonstrated [10,11] and the WHO has recommended their use in cervical cancer screening, as it believes they will lead to greater acceptance of and participation in screening programmes [12-14]. Thus, in line with European recommendations, in 2019 the Spanish Ministry of Health determined [15] the need to implement population-based cervical cancer screening programmes throughout the country and established HPV testing as a screening test for women aged 35 to 65. The country’s various regions are assessing the use and implementation of self-sampling devices.

Therefore, given that high target population participation and coverage rates are necessary to achieve the desired programme effectiveness, strategies adapted to the specific sociological, cultural, and healthcare characteristics of each region and community must be designed prior to the implementation of a population-based programme [16]. The characteristics of the population and the programme (such as the type of screening test used, the way the target population is invited, uptake, or information campaigns) are a key factor in the results [17]. For this reason, to reach the established goals, it is fundamental to study the population where the programme is to be implemented in order to find out their preferences, needs, barriers, or insecurities regarding different models of invitation, uptake, information, screening tests, etc., thus enabling equitable access to the programme [18].

As regards selecting strategies to optimise screening programmes, the incorporation of new technologies (mobile applications or social networks) to inform, invite, and ensure uptake of the target population is an opportunity for improvement in new screening models. Some of these tools are already being used and tested to determine how they could improve participation in population-based cancer screening programmes [19,20], and they must be assessed when implementing a new programme.

In this line, and given that primary screening test uptake is of the utmost importance in achieving high participation rates, the assessment of new self-sampling devices is essential. Even though numerous studies show that they are greatly accepted by women [21,22] and they are included in the WHO Consolidated Guideline on Self-Care Interventions for Health [23], there are limited studies assessing the type of device that women prefer from the wide range available on the market [24]. However, their format and mode of use can be a critical factor in population participation, and recently published studies have highlighted the need for additional research examining women’s preferences [24]. This will be essential to reach the WHO’s target that 70% of women should undergo cervical cancer screening by 2030 [25].

As a population-based cervical cancer programme will soon be implemented in the Valencia Region, the purpose of this study was to find out the expectations, preferences, and difficulties perceived by women as regards different ways of inviting them to participate in the prevention programme, uptake and self-sampling devices for HPV testing, with the aim of detecting strategies to achieve high participation rates in this region’s population-based cervical cancer prevention programme.

## Materials and Methods

The study was based on qualitative research techniques.

### Ethics statement

All methods of the study were performed in accordance with the Declaration of Helsinki and was approved by the It was approved by the Committee of Ethics and Research Integrity of Miguel Hernández University of Elche (reference no. AUT.DPS.JMS.01.21, date 30/09/2021).

Participants were informed and subsequently signed a consent form to participate in the study and agreed to the sessions being recorded.

### Design and Participants

We worked with a total of 29 women. The following inclusion criteria were used for non-random selection: accept to participate, be aged between 35 and 65, live in the Region of Valencia, and represent different ethnicities. We created mirror groups by working in parallel with 4 sets of 7–8 women who had a similar profile in terms of age, educational level, and professional activity.

The women were selected using the snowball technique, ensuring that working and discussion groups were sufficiently representative in terms of age, education, professional activity and ethnicity.

Sessions were conducted with the four groups simultaneously, lasted 120 minutes each, and were recorded with the consent of the participants. The anonymity and confidentiality of the recorded material was guaranteed. The process was led by researchers with extensive experience in conducting qualitative studies.

### Interventions and content

The <u>first phase</u> of the session involved an individual interview with each woman, where they anonymously completed a **questionnaire of semi-structured questions** in writing using a workbook. This questionnaire was carried out individually and anonymously to prevent the participants from influencing each other. In the <u>second phase</u>, **group interviews** were used to confirm trends or reveal any possible points of interest that had not been addressed.

During the <u>first phase</u>, the women were presented with a draft programme information leaflet regarding the new cervical cancer screening programme, which included the use of self-sampling devices as a screening method for women aged 35 to 65. These devices would be received by post and returned in health centres. Women responded to questions about the content of information materials, their preferred information channels, and details of the screening model and its circuits at each stage (method of invitation, acceptance and use of the self-sampling device, method of communicating results, etc.). Multiple-choice questions were used for this purpose. They also answered questions about their previous participation in the opportunistic cervical cancer screening programme or whether they were aware of other screening programmes.

Subsequently, the self-sampling devices were distributed to the women in a sequence and they individually assessed each one without influencing the views of the other women in the group. Manufacturer instructions on how to use the device were provided. Five tests were assessed in this study (figure 1): (Device A) FLOQSwab^®^ 552C.80 (Copan Diagnostica Inc.), (Device B) Evalyn^®^ Brush (Rovers Medical), (Device C) Aptima Multitest Swab^®^ (Hologic), (Device D) IUNETEST^®^ (Self Test Technologies, S.L), and (Device E) Qvintip^®^ (Aprovix). They were selected based on their previous use in screening programmes and their ease of use. When the devices were distributed, the researchers provided usage instructions and the participants then answered two questions regarding whether they thought they were easy to use and if they would use them at home, plus an open-ended question to explain their opinions. The devices were presented to the 4 groups in a random order. After the individual assessment, the devices were compared and the women selected which one they thought was the easiest to use, which one they felt most comfortable or confident using, which one they considered the best and worst, and which three devices they would prefer to use.

**Fig 1.**
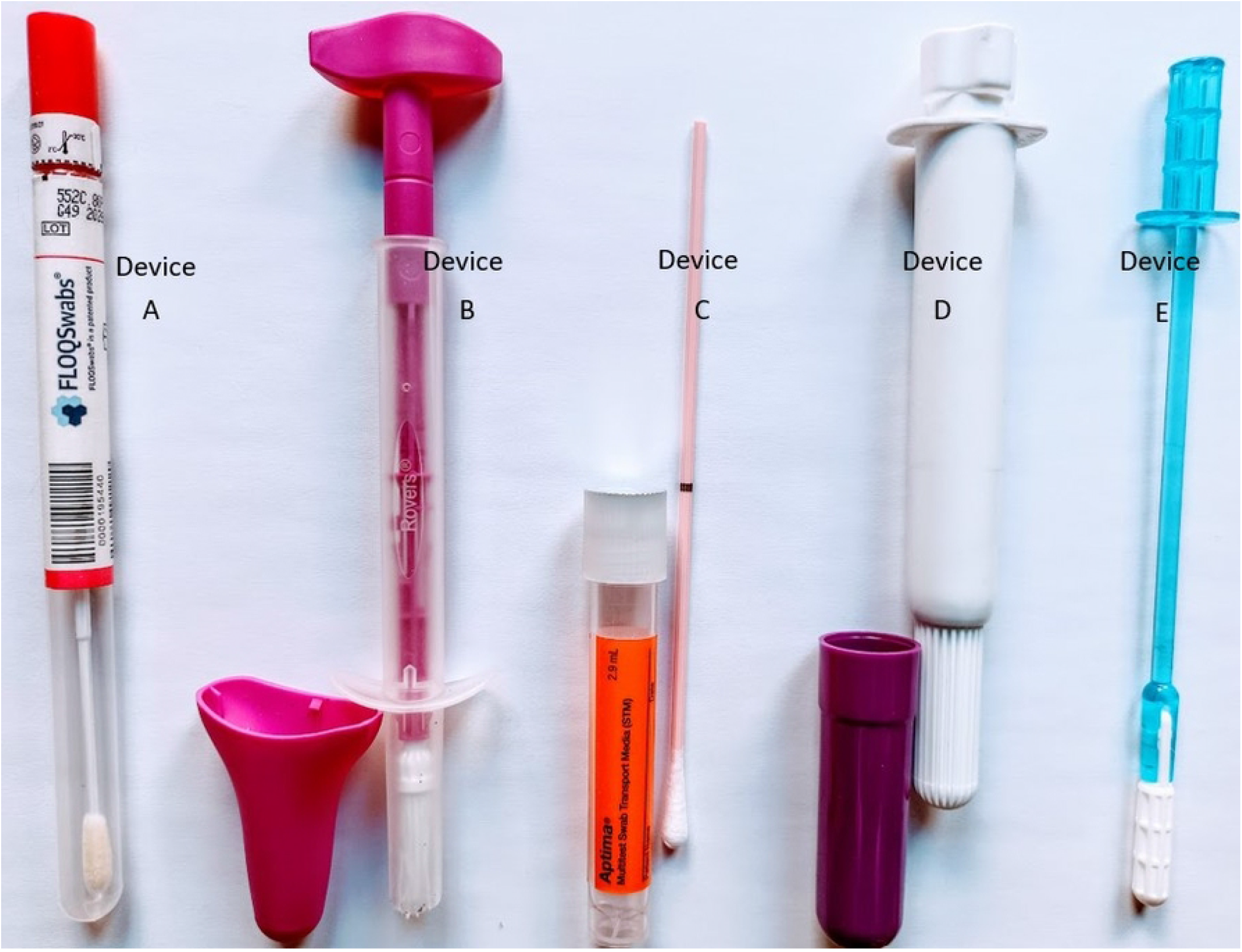
Self sampling devices assessed. (Device A) FLOQSwabs is a plastic swab with a rounded tip made of nylon fibers, which must be removed from the tube containing it by rotation, inserted into the vagina up to the indicative line and reintroduced into the tube; (Device B) EvalynBrush is a plastic device, with a brush at one end and a plunger at the other end, with a cap. It must be inserted up to the side wings, rotate the plunger 5 times and put the lid on; (Device C) Aptima Multitest Swab consists of a cardboard swab with a rounded cotton tip and a tube with a liquid medium. After inserting the swab into the vagina up to the indicative line, it must be inserted into the liquid medium and broken to leave the tip inside; (Device D) IUNETEST is a plastic tube with a cap at one end and a rotary plunger at the other. To use it, it must be inserted, the plunger pushed and rotated, and finally the lid is placed; (Device E) Qvintip contains a test tube and a wand with a hooked tip at one end. For use, after inserting and rotating the wand in the vagina, the plastic tip must be inserted into the tube, resting on its walls to unhook it of the wand and leave it inside.

<u>During the group interview</u>, a structured discussion was opened in which the women talked about the new screening programme, the content of the information leaflet, and the self-sampling devices. Data were triangulated to analyse similarities and differences between the groups. Interviews were ended once a sufficient level of information saturation had been reached.

### Analysis

A descriptive analysis was created showing frequencies and percentages relative to the different options provided in the closed-ended questions. For the open-ended questions, the session transcripts were used to identify and classify the different ideas into mutually exclusive categories. The number of different ideas was counted, as well as the number of times each of these ideas was repeated, as a measure of contribution intensity. Two researchers assigned the ideas to categories. A third researcher was consulted if they were unsure or unable to reach an agreement, and a consensus was reached regarding the assigned category for all ideas.

When analysing the group interview results, the spontaneity of the women’s ideas was taken into account. This refers to the number of women who independently proposed the same idea. The data was then triangulated to analyse similarities and differences between participants and groups of participants.

## Results

A total of 29 women participated in the study. In terms of educational level, 44.8% (13 women) had completed university studies, 27.6% (n=8) had completed vocational training studies, and the rest had completed compulsory secondary education. The average age was around 50. A total of 96.6% of women (n=28) had had a cervical cytology test, and 86.2% (n=25) had had a cervical cytology test every 3 years or less. In addition, 79.3% (n=23) said they were aware of other cancer prevention campaigns (breast and colon).

### CCSP assessments

When assessing different aspects of the programme (figure 2), it was found that women preferred to <u>receive information</u> on the programme from health centres (86.2%), from breast cancer prevention units (75.9%), from women’s associations (69%), by post (51.7%), and by SMS (44.8%), or through email, citizens’ associations, and work centres (34.5% overall). In relation to the <u>tools for contacting</u> the population, the women preferred to be invited to participate in the programme by SMS, selected by 58.6%, or by post with 48.3%, while the mobile application of the Regional Ministry of Health was accepted by 24.1% of women. Similarly, the majority chose to <u>receive test results</u> by SMS (65.5%), while 37.9% preferred to receive results by post and 20.7% through the mobile application. Once the sample had been collected, they preferred to <u>return their self-sampling devices</u> in health centres (86.2%), as opposed to pharmacies (17.2%) or hospitals (3.4%). In relation to <u>screening by self-sampling</u>, 96.4% stated that they would take part if this screening method were used.

**Fig 2.**
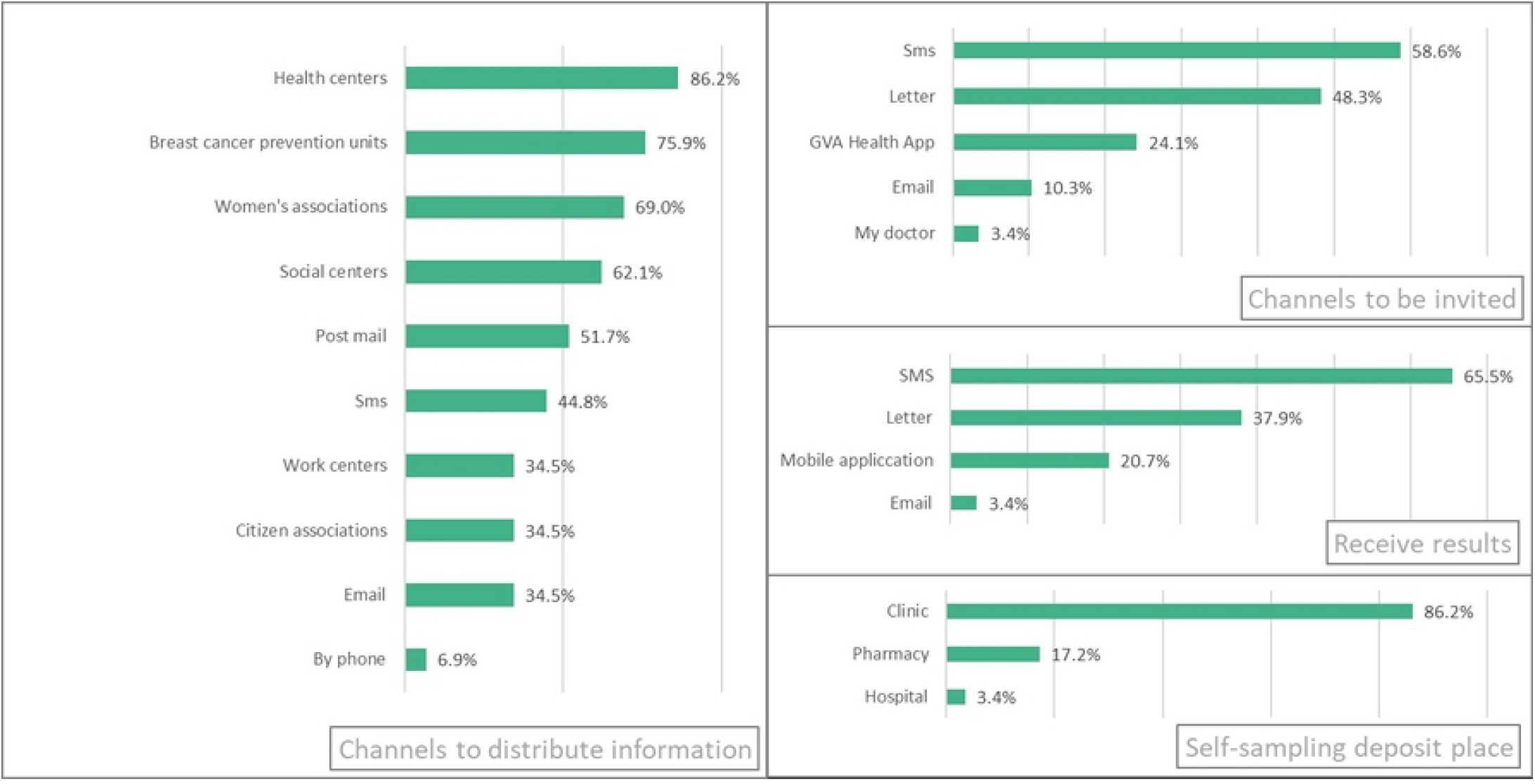
Preferences relating to different general aspects of the screening program.

### Individual device assessment

Overall, 93.1% of women considered that Device A was easy to use, 79.3% Device D, 69% Device B and Device C, and 62.1% Device E. The most widely accepted devices for home use were Device A and Device D, both with 75.9% acceptance, followed by Device B (72.4%), Device E (48.3%), and Device C (37.9%). The results are shown in figure 3.

**Fig 3.**
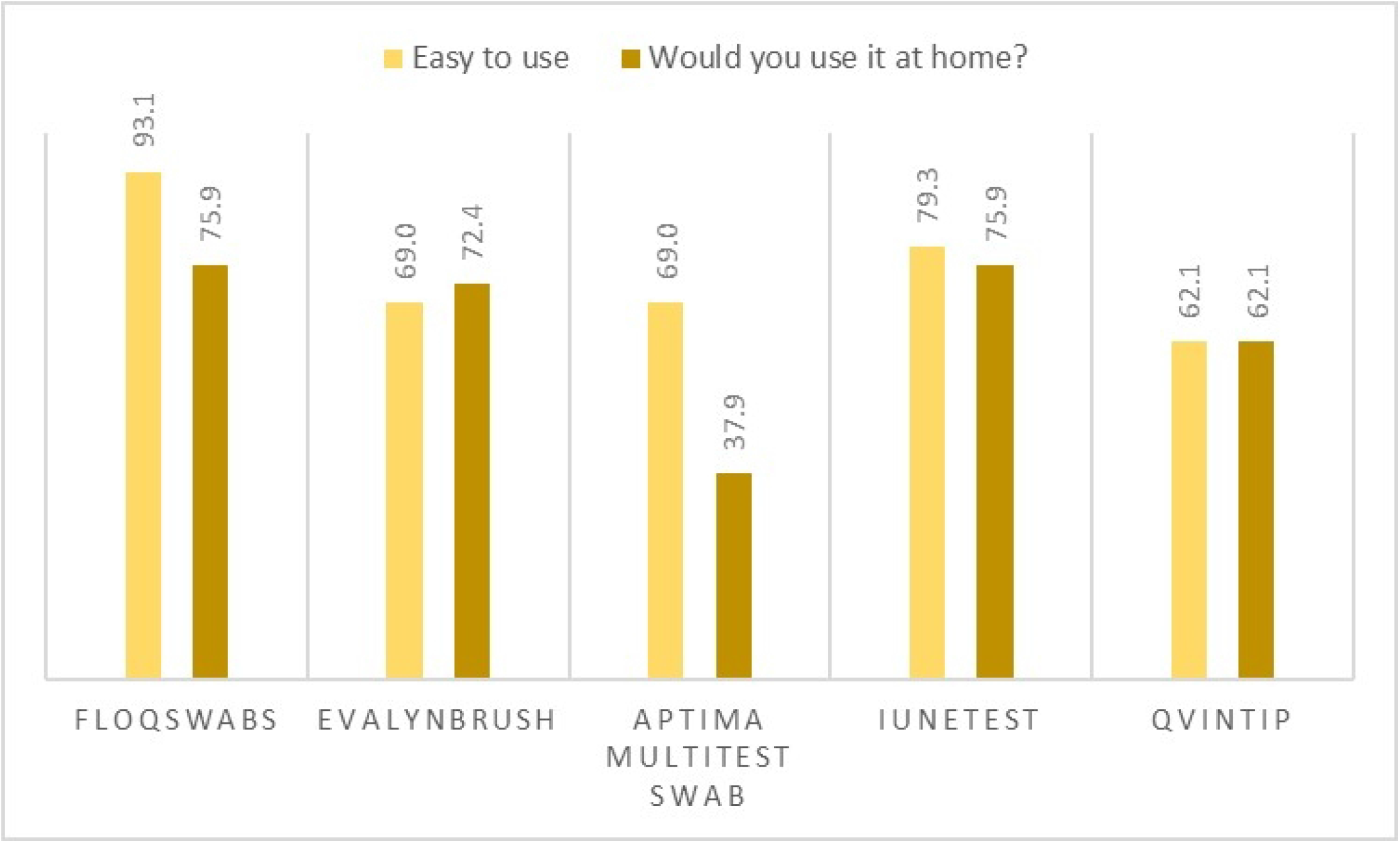
Results of the individualized assessment of devices according to ease of use and their acceptance for use at home.

Various **positive aspects** were highlighted in the open-ended question (data shown in table 1). One example is <u>simplicity and ease of use</u>, which was mentioned by 15 women in relation to Device A and 13 women in relation to Device D. Another positive aspect pointed out by the women was that <u>the device was not perceived to be invasive</u>, as indicated by 4 women in relation to Device A, Device C, and Device D. The <u>attractive design</u> of Device B was mentioned by 6 women. In addition, 4 women considered that the <u>familiar</u> (tampon-like) appearance of Device D was positive. The participants identified the highest number of different positive aspects in relation to Device A and Device B, with a total of 11 each. The total number of times that these positive aspects were mentioned overall was highest in relation to Device A, with a total of 30. The most frequently indicated **negative aspects** were <u>difficulty of use and operation</u>, which were mentioned by 9 women in relation to Device C and 7 in relation to Device E. <u>Discomfort</u> was another aspect mentioned by 5 women in relation to Device D and Device E, and by 4 women in relation to Device B. A total of 7 women highlighted the high risk of <u>sample contamination</u> with Device E due to the usage method. <u>Fragility</u> was rated as a negative aspect by 5 women in relation to Device A and by 3 in relation to Device C. The women identified the highest number of different negative aspects in relation to Device E (10), Device B (8), and Device C (8). The total number of times that these negative aspects elements were mentioned overall was highest in relation to Device C (26), Device E (22), and Device B (18).

**Table 1.** Positive and negative evaluations of the self-sampling devices collected from the open-ended question. Due to the great diversity of aspects mentioned, this table includes the most relevant, having left some aspects out of the table, which is why the total number of elements does not correspond in many cases to the total number of the list.

|  | FLOQSwabs | EvalynBrush | Aptima Multitest Swab | IUNETEST | Qvintip |
| --- | --- | --- | --- | --- | --- |
| **Positive aspects** |  |  |  |  |  |
| Simplicity of the device, ease of use | 15 | 4 | 5 | 13 | 5 |
| non-invasive device | 4 | 3 | 4 | 4 | 2 |
| Safe, inspires confidence | 2 | 3 | 1 | - | 3 |
| Family mechanism (similar to a tampon) | - | - | - | 4 | 1 |
| Mark indicating the depth of penetration | 2 | - | 1 | - | - |
| Attractive design | - | 6 | - | - | - |
| **Total (Different items/totals)** | 11/30 | 11/23 | 8/15 | 9/27 | 5/12 |
| **Negative aspects** |  |  |  |  |  |
| Use and complex handling | 4 | 4 | 9 | 3 | 7 |
| Uncomfortable | 2 | 4 | - | 5 | 5 |
| Sudden steps, which requires strength | - | 4 | 2 | - | 1 |
| High risk of sample contamination | - | 1 | 3 | - | 7 |
| Fragile | 5 | - | 3 | - | - |
| Dangerous | - | - | 4 | - | - |
| Unhygienic | - | - | 3 | - | - |
| Low trust, unsure | - | - | - | - | 4 |
| **Total (Different items/totals)** | 5/11 | 8/18 | 8/26 | 5/9 | 10/22 |

### Comparative device assessment

The most voted devices were as follows: for ease of use, Device A (52.38%) and Device D (28.57%); for feeling of confidence transmitted, Device B (51.72%) and Device A (24.14%); for comfort of use, Device B (35.71%) and Device A (25.0%). When asked to select the device they liked the most, Device B (31.03%) and Device A (27.59%) received the most votes, and when asked to select the one they liked the least, Device C (28.57%), Device E (25.0%), and Device B (21.43%) received the most votes. The results are shown in figure 4. Finally, when participants were asked to select the three self-sampling devices they would use at home, Device A (22), Device B (20), and Device D (18) received the most votes, while Device C and Device E were selected by 13 and 11 women, respectively (results not shown in tables).

**Fig 4.**
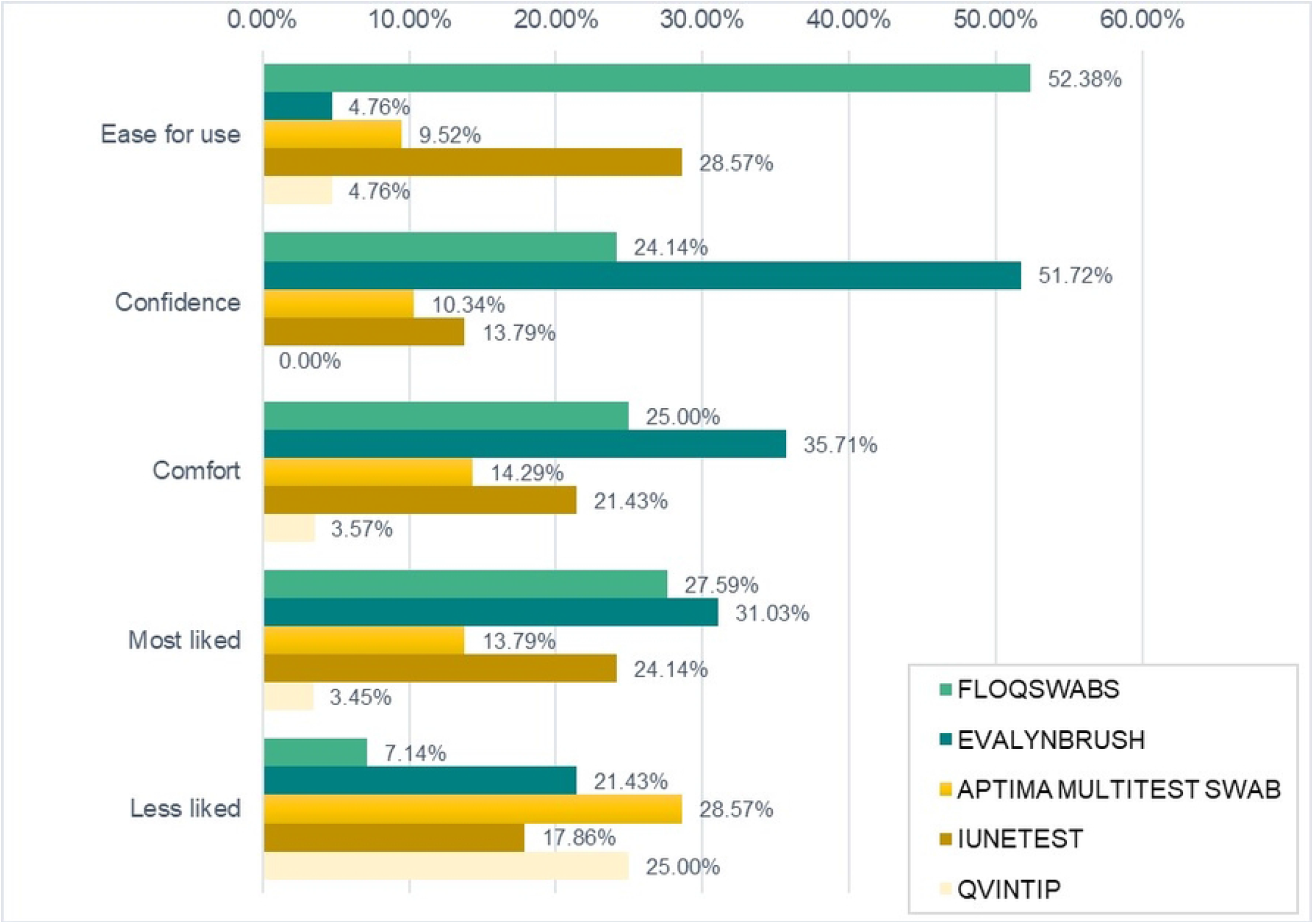
Results of the selection of a device for its greater ease, reliability, comfort and more and less appreciated.

### Self-sampling assessment synthesis

Thus, Device B is preferred by some women, and Device A is accepted by the greatest number of women. Simplicity and ease of use are the key features highlighted in relation to Device A, the device with the highest number of votes. The attractive design of Device B was mentioned as a positive aspect and, although it was some women’s preferred option, others considered that it was uncomfortable to use. Device C and Device E were accepted by the lowest number of women. The self-sampling process of devices with lower levels of acceptance included intermediate steps or steps perceived as complicated due to their rough or dangerous nature. Examples are the need to insert the sample into a liquid medium in the case of Device C, or the painstaking process of removing the crimped tip from the sampler stick in the case of Device E. Although Device D was not singled out due to any negative aspect in particular, it did not score well in the comparative assessment.

### Group session assessment

Finally, ideas that came up in the group session revealed many positive attitudes towards a new screening programme based on self-sampling devices (n=9). The existence of another screening programme based on a self-sampling device (faecal occult blood screening devices used in the colorectal cancer prevention programme) is seen as an opportunity (n=3). Some verbatim examples are as follows: “*If you can take the self-sample test for the colon cancer prevention programme, you can do this one, too*”, “*I think it’s great that everyone takes their own sample*”, “…*we’re already familiar with the colon cancer programme*…*or know someone that has taken part*… *I think it will be much easier to implement the cervical cancer prevention programme*”. In addition, home testing is considered positive due to the speed and practicality (n=1) of this method, as well as the possibility of including more vulnerable groups (n=2). *“People of different ethnicities or races will have a better chance of being screened”, “it can reach more people and free up in-person appointments”, “you don’t have to wait, because sometimes you can’t be bothered to go to the gynaecologist to get it done*”. They believed that this programme will be accepted, on the one hand, due to women’s increased awareness of screening (n=2) and, on the other hand, because they consider that people will eventually adapt to the new measures (n=1).

Ideas were provided for improving the content of the message in information leaflets or possible campaigns, showing the need for a stronger message (n=6): “include information on mortality”, “…more forceful messaging”.

Perceived fears centred on uncertainty as regards the reliability of self-sampling (n=4): “*maybe we won’t do the test correctly and the results aren’t right”, “questions about precision: how many centimetres it should be, counting the turns, etc*.*”, “uncertainty of not going to a clinic/hospital and not noticing any other illnesses or mistakes with self-sampling*”. The feeling of a lack of protection for certain age groups or groups that might not be able to use the self-sampling device due to a lack of knowledge about their own bodies also generated uncertainty. Here are some verbatim statements: “*feeling of lack of protection and information for older people”, “due to their age, they prefer traditional screening”, or “many women do not have sufficient knowledge of their own bodies to do this themselves*”. The statements grouped by theme are shown in table 2.

**Table 2.** Verbatim statements provided in the group session to assess the cervical cancer screening programme and self-sampling, grouped by category. The figures in brackets represent the number of times the message was mentioned by women.

| Categories | Verbatim statements |
| --- | --- |
| Positive attitude to the new self-sampling programme (9) | Previous positive experience with the colon test. *"It's something that gets sent to your home, that you have to do, you read the instructions, you follow them, you take the sample, you take it to the health centre and then they send you the letter" "You're in perfect health" or "no, bad luck". "If you are not well, the doctor calls you. If you can take the self-sample test for the colon cancer prevention programme, you can do this one, too. I think it's great that everyone takes their own sample"* (3) |
|  | People of different ethnicities or races will have a better chance of being screened. It can reach more people and free up in-person appointments. It can also be useful to monitor how often you have had a Pap smear. (2) |
|  | Increased awareness of HPV (2). *"I think women are more aware that we could get cancer or the human papilloma virus. So, I get the impression that we won't think it's strange to get the device and take the self-sample at home". "I think it will be easy and quick to set up for all the reasons that the others are saying. We're already familiar with the colon cancer programme or know someone that has taken part. I think it will be much easier to implement the cervical cancer prevention programme"* |
|  | It is practical. *"You don't have to wait, because sometimes you can't be bothered to go to the gynaecologist to get it done"* (1) |
|  | A matter of time and practice to adapt to new measures (1) |
| Content of the message in information materials (6) | More forceful messaging (2) |
|  | Messages must be very forceful to ensure that the programme doesn't generate doubts (1) |
|  | Mention the reasons for the screening programme with health statistics: mortality, incidence (2) |
|  | "For example, just because you don't have any symptoms doesn't mean you are unhealthy, you could have HPV and develop cervical cancer in the future" (1) |
| Lack of information on the programme itself (5) | More information if restricted to cervical cancer or other diseases (2) |
|  | The only doubt is where to drop it off at the health centre (1) |
|  | It may give rise to doubts (1) |
|  | Provide information on the possible infections and discomfort that it can cause (1) |
| Reliability of self-sampling (4) | Maybe we won't do the test correctly and the results aren't right (1) |
|  | Uncertainty about whether you have done it right (1) |
|  | Questions about precision: how many centimetres it should be, counting the turns, etc. (1) |
|  | Uncertainty of not going to a clinic/hospital and not noticing any other illnesses or mistakes with self-sampling (1) |
| Feeling of a lack of protection due to insufficient knowledge of the body or not being in the age range of the programme (5) | It starts at 25 — Younger women are vaccinated against HPV (1) |
|  | Feeling of lack of protection and information for older people (1) |
|  | Due to their age, they prefer traditional screening (1) |
|  | The age from which it is necessary (1) |
|  | Knowledge of the body: "Many women do not have sufficient knowledge of their own bodies to do this themselves" (1) |

## Discussion

The results of this study show numerous opportunities to implement a new population-based cervical cancer screening programme.

On the one hand, the use of self-sampling devices is seen as an opportunity as, according to numerous publications [24,26,27] this method is highly accepted by women regardless of the device used. This can also be seen in the results of our study. It is demonstrated by both the high acceptance (96.4% of women) of its use as a screening test and the perceived advantages of use (ease of use, uptake of certain groups, etc.).

On the other hand, the existence of established population-based programmes, some of which also include the use of self-sampling tests such as the faecal occult blood test for colorectal cancer screening, could facilitate the acceptance of self-sampling devices for HPV testing, as highlighted by the women in this study.

Similarly, the high acceptance of SMS messaging or health system mobile applications as a means of communication, which could be because the COVID-19 pandemic consolidated the use of new technologies in the healthcare environment, could also facilitate the implementation of the programme as well as greater uptake and participation [28,29].

Despite the high acceptance of self-sampling, many women are uncertain of their ability to perform the sampling procedure correctly, as has also been pointed out in numerous studies [30-33], or fear that other diseases may not be caught if they do not attend an in-person visit, as per usual practice in opportunistic screening. In addition, while self-sampling may enable vulnerable groups who may find it more difficult to attend an in-person visit to participate, as some studies on the use of self-sampling devices in disadvantaged populations have shown [34], it may also leave certain population groups unprotected. Therefore, to improve women’s confidence in self-sampling, public awareness campaigns should include data showing that most women can successfully obtain an adequate sample [18,31,35,36]. Furthermore, information campaigns should also address the lack of protection felt by women in the absence of an in-person visit. To fight against inequality, adapted materials must be created in different languages that are easily understandable for different socio-cultural levels, as pointed out in the WHO Guideline on self-care interventions for health and well-being [17].

As regards the content of these information campaigns and materials, the women in the study believe that it is important to highlight the serious nature of the disease in order to raise public awareness. In this line, the literature indicates that increased risk perception leads to increased participation in screening programmes, and also to increased acceptance of the use of self-sampling devices in the case of cervical cancer [37-38]. Furthermore, appropriate public awareness and information campaigns and educational actions to raise awareness of the importance and impact of cervical cancer prevention have shown to be effective in increasing the acceptance of self-sampling [34,39].

The results of this study reveal differences in the degree of acceptance of the different self-sampling models, as was also observed in several studies [31,40,41]. The perceived difficulty when using the device, a feature mentioned in relation to the Qvintip or Aptima Multitest Swab devices, appears to have a significant effect on preference as women liked these [41]two devices the least. An attractive design that is specifically conceived for self-sampling, as described previously, is one of the main features of self-sampling devices that seem to transmit confidence due to their colour or appearance. This quality makes the Evalyn Brush one of the most highly rated devices for some women. However, simplicity is the most important feature of the device accepted by the largest number of women. Other comparative studies [42] also show that women rate this aspect positively in their choice of self-sampling device. Thus, the swab-type device (FLOQSwab) is accepted by the largest number of women, as was observed by Nishimura et al [24]. Although there are not many studies comparing acceptance of different self-sampling devices, there are studies comparing swab and tampon devices which show that the former is more widely accepted, as in this study [43].

Some studies do not agree with the results obtained in this study, such as the one carried out by DiGennaro et al, which observed no differences in the degree of acceptance with different types of self-sampling device [44]. This may be because preferences can be influenced by population characteristics (cultural, religious and socio-economic) that determine the degree of acceptance of different devices. Therefore, it may be useful to conduct a pilot study prior to full-scale implementation of the programme, as suggested by Arbyn et al [45].

### Limitations

Nearly half of the participants had completed university studies, which is an over-representation of this group. Another limitation is that the women were able to read instructions on how each of the devices work and operate them, but not to actually use them to take a sample when carrying out their assessment.

The lack of published studies comparing different self-sampling devices, as well as the lack of homogeneity of these devices, makes it difficult to compare results.

## Conclusions

The results of this study provide further insight into preferences regarding self-sampling formats for cervical cancer screening. These results can aid decision-making to implement or improve cervical cancer screening programmes by selecting models that are more widely accepted in order to achieve higher participation rates.

The use of new technologies (SMS messaging or specific mobile applications), probably driven by the COVID-19 pandemic, stands out as an effective strategy for reaching out to the population in screening programmes.

While self-sampling presents an opportunity for the implementation of population-based screening, a screening model based on self-sampling must go hand in hand with powerful and adapted information, education, and public awareness campaigns to build the most vulnerable groups’ confidence in self-sampling in order to achieve high participation rates in all population groups and thereby reduce inequalities. Thus, as high participation is a key objective of the programme, appropriate campaigns and educational interventions to raise awareness of the importance and impact of cervical cancer prevention have shown to be effective in increasing the acceptance of self-sampling [34,39].

It can be concluded that simplicity is an essential aspect when selecting the self-sampling device. Additionally, given the diverse range of opinions on the different self-sampling models, preferences should be taken into account to select a device that allows for the greatest level of programme participation.

Although the use of self-sampling devices has been studied primarily to increase uptake by non-responding women [46-52] or specifically targeted at disadvantaged groups due to their socio-economic status or health conditions [53] [54-58], it is already being used as a screening method for the general population in some countries such as the Netherlands [59] and Australia [60]. As recent studies point out [61], although self-sampling may entail a potential loss in sensitivity, this is likely to be offset by the improved effectiveness of the programme resulting from increased acceptance, and it is a more equitable option for women.

Lastly, within this new self-sampling model, we must not forget essential aspects in the implementation of population-based screening, such as the use of validated tests [62-63] and the importance of evaluating the results or continuity strategy for follow-up in the case of women with a positive high-risk HPV result.

## Data Availability

All methods of the study were performed in accordance with the Declaration of Helsinki and was approved by the Responsible Research Office of Miguel Hernández University (reference no. AUT.DPS.JMS.01.21, date 30/09/2021). Participants were informed and subsequently signed a consent form to participate in the study and agreed to the sessions being recorded. All methods were performed in accordance with the Declaration of Helsinki relevant guidelines and regulations.

## List of abbreviations

HPV: Human Papillomavirus
CCSP: Population-based cervical cancer screening programmes
IARC: International Agency for Research on Cancer

## Acknowledgements

The authors would like to thank all the women that participated in the study for their time and contributions.

## Notes

### Competing Interest Statement

The authors have declared no competing interest.

### Funding Statement

This research did not receive any specific grant from funding agencies in the public, commercial, or not-for-profit sectors.

